# Adiposity and the Prevalence, Incidence and Distribution of Refractive Errors in Urban Vietnamese Children

**DOI:** 10.64898/2026.09.17.26363286

**Authors:** Nhan Thi Ho, Khoi Pham Minh Tran

**Affiliations:** Research Management Department, Vinmec International Hospital, Hanoi, Vietnam; Pediatric Department, Vinmec Times City, Vinmec International Hospital, Hanoi, Vietnam; College of Health Sciences, VinUniversity, Hanoi, Vietnam

## Abstract

**Purpose:** To examine the association between BMI-for-age z-score (BAZ) and BMI status with refractive error prevalence, incidence, subtypes, and quantitative refraction in Vietnamese schoolchildren.

**Methods:** We retrospectively analyzed body mass index and refractive error data from annual school health checks in three Vietnamese cities, 2022 to 2024. The main analysis set was restricted to children age 6 and older (49,415 examinations, 31,522 children). Refractive error subtypes and quantitative refraction came from a Hanoi autorefractor sub-cohort (13,059 examinations). We used sex-stratified Poisson models with generalized estimating equations, adjusted for age, city, and year, plus linear mixed models for quantitative outcomes.

**Results:** Any refractive error followed a U shaped curve across BAZ in both sexes, significant in girls (prevalence ratio 1.01 per 1-SD BAZ, p = 0.027) but not boys (p = 0.099). Obesity was associated with higher any refractive error prevalence in girls (adjusted prevalence ratio 1.05, p = 0.017). Astigmatism was associated with BAZ in both sexes (p < 0.001), anisometropia in boys only (p < 0.001), with no association for myopia or hyperopia. Spherical equivalent shifted slightly toward hyperopia, not myopia, with higher BAZ. A supplementary check over 1 to 2 years found no evidence that prior BAZ predicted new refractive error onset (prevalence ratio 1.02, p > 0.6 in both sexes), and the nonlinear pattern did not reappear.

**Conclusions:** Adiposity relates cross-sectionally to refractive error in a nonlinear, sex specific way concentrated in astigmatism and anisometropia rather than myopia, though this did not hold up short term.

## INTRODUCTION

Over the last decades, childhood refractive errors, especially myopia has become one of the most common chronic conditions in pediatric eye care in Vietnam, Southeast Asia and worldwide ^1,2^. Overweight and obesity have also risen sharply worldwide, with the most recent pooled analysis of population based measurement studies estimating that tens of millions of school age children now live with obesity across income settings ^3^. Because both trends track the same shift toward urban, screen heavy, indoor oriented childhoods, researchers have increasingly asked whether adiposity itself, not just the environment that produces it, contributes to refractive error.

The evidence linking body mass index (BMI) to refractive error in children is substantial in volume but inconsistent in shape. A 2025 systematic review and meta-analysis pooling data across the pediatric literature reported significant positive associations between anthropometric measures and myopia, with pooled odds ratios of 1.27 for overweight and 1.25 for obesity, though heterogeneity across studies was high ^4^. The largest single study to date, a nationwide Israeli cohort of 1.36 million adolescents, found a J shaped pattern in which both underweight and high body mass index were associated with myopia, with larger effect sizes in males ^5^.

Findings from Korea’s National Health and Nutrition Examination Survey showed obesity associated with a 3.77 fold higher odds of high myopia, with the strongest effects in girls, and a predicted high myopia prevalence rising from 5.3% in underweight adolescents to 20.6% in those with obesity ^6^. A nationwide Chinese study similarly found stronger overweight and obesity associations with myopia in girls age 6 to 11 ^7^, while another Chinese sample described an inverse L shaped rather than linear dose response curve ^8^. Obesity has also been linked to roughly two fold higher odds of astigmatism in at least one large pediatric study, though hyperopia has been studied far less and the evidence there remains sparse ^9^. In the United States, NHANES based analyses have found positive associations between adiposity indices and myopia ^10^, but a Mendelian randomization analysis using much of the same evidence base did not support a causal genetic effect, which weakens confidence in a direct biological pathway ^11^.

This current literature has three limitations. First, it is almost entirely cross sectional and cannot separate confounding by outdoor time, near work, parental myopia, and socioeconomic status from a true adiposity effect ^6,12^. Second, the reported shape of the association varies by population, ranging from linear to J shaped to inverse L shaped, so a single pooled odds ratio may obscure real heterogeneity by age and sex. Third, and most relevant here, Southeast Asia is represented almost entirely by Korea and China, with a small Indonesian study finding no association between BMI status and refractive error ^13^, and no study has directly tested the obesity to refractive error association in a Vietnamese population, despite Vietnam’s documented refractive error burden ^2^.

This study uses repeated school health examinations from a large, multi city Vietnamese cohort to address the last two gaps in literature mentioned above. We estimate the cross sectional association between BMI-for-age z-score and BMI status category and the prevalence of any refractive error, refractive error subtypes, and quantitative spherical equivalent and cylinder, allowing separately for sex and for a nonlinear BMI-for-age relationship given the mixed shapes reported elsewhere in the literature. We also use consecutive annual visits to test whether the cross-sectional association predicts new onset of refractive error.

## METHODS

### Study design and population

This study conducted a retrospective analysis of de-identified data from annual school health examinations of children aged 3 to 18 years between 2022 and 2024 in three major Vietnamese cities: Hanoi, Ho Chi Minh City, and Hai Phong. Records with complete BMI and refractive-error information at the same visit were included in the analysis.

### Anthropometric exposure

BMI-for-age z-scores (BAZ) were calculated separately for boys and girls and categorized as four categories (thinness, normal BMI, overweight, obesity) using LMS method and 2007 World Health Organization (WHO) growth references ^14,15^. Absolute BAZ of ≥ 6 were excluded. BAZ was analyzed primarily as a continuous exposure, with associations expressed per 1-SD increase and secondarily as four categories.

### Refractive-error outcomes

The primary outcome was any refractive error based on the refractive-error classification recorded in the school screening dataset. Secondary outcomes available in the Hanoi autorefractor subgroup included myopia, moderate or high myopia, hyperopia, astigmatism, and anisometropia as well as quantitative refractive outcomes. Spherical equivalent (SE) for the right and left eyes was analyzed as a continuous measure in diopters. Absolute cylinder power was analyzed as a continuous measure of astigmatic refractive magnitude. More negative SE values indicate a more myopic refractive state.

### Statistical analysis

Crude prevalence estimates were presented with 95% Wilson confidence intervals (95%CI) ^16^. The primary association between BAZ and any refractive error was estimated by prevalence ratios using a modified Poisson regression model with a log link fitted using generalized estimating equations (GEE) ^17^, exchangeable correlation, and robust sandwich variance to account for correlation among repeated examinations ^18^.

All models were fit separately for boys and girls. The primary model adjusted for age (natural spline), city, and examination year, and a nonlinear model added a spline for BAZ, tested with a joint Wald test. Adjusted prevalence curves were generated across the observed BAZ range, holding other covariates at sex-specific values.

BMI status categories used normal weight as reference. City-specific analyses and BAZ by city interactions were exploratory. Secondary outcomes used the same GEE framework. Any refractive error was the primary outcome and received no multiplicity adjustment. The five secondary subtypes were Benjamini-Hochberg false discovery rate adjusted, within each sex ^19^.

For quantitative refractive measures in the Hanoi autorefractor subgroup, linear mixed effects models with a random intercept for child adjusted for BAZ, age, and year, with natural splines checking nonlinearity, fit with lme4 ^20^.

Sensitivity analyses included the full age 3 to 18 range, restriction to age 6 and older, raw BMI instead of BAZ, city-specific analyses, and exclusion of current spectacle wearers.

### Longitudinal replication check

Because the primary analysis used repeated cross-sectional examinations, it could not establish temporal order. As a supplementary check, we built consecutive visit pairs within each child, using the same two analysis sets, so BAZ at the earlier visit predicted status at the later visit among children free of the outcome earlier. The follow-up time of most pairs were 1 year, with a few spanning 2 years which is a limitation of our data. Incident any refractive error was modeled with the same sex-stratified GEE framework, adjusted for age (spline), city, and year, with linear and nonlinear BAZ terms. The same approach covered incident astigmatism, anisometropia, myopia, moderate or high myopia, and hyperopia in the Hanoi subgroup, with Benjamini-Hochberg correction across these five outcomes within each sex. We first tabulated children at risk, incident cases, and person-years for each outcome, since several subtypes had too few events for meaningful estimation. As a second check, each child’s BAZ averaged across all available visits predicted whether a child free of any refractive error at their first visit had developed it by their last visit.

All statistical tests were two-sided. A P value <0.05 was considered statistically significant for the prespecified primary analysis. Statistical analyses were performed using R version 4.5.1 ^21^.

## RESULTS

### Study population and participant characteristics

The study dataset included 68,097 examinations with complete BMI and refractive-error information at the same visit. The main analysis set included 49,415 examinations from 31,522 children (25,583 examinations in boys, 23,832 in girls, median 2 examinations per child). The Hanoi autorefractor subgroup itself contributed 13,059 examinations for the refractive error subtype and quantitative analyses (6,978 boys, 6,081 girls)

The distribution of BAZ varied substantially with age and differed by sex (**Figure 1**). In the main analysis set, mean age of boys was 11.0 years, and of girls was 11.2 years (**Table 1**). Age distributions differed significantly across BMI-for-age categories in both sexes (p<0.001).

**Figure 1.**
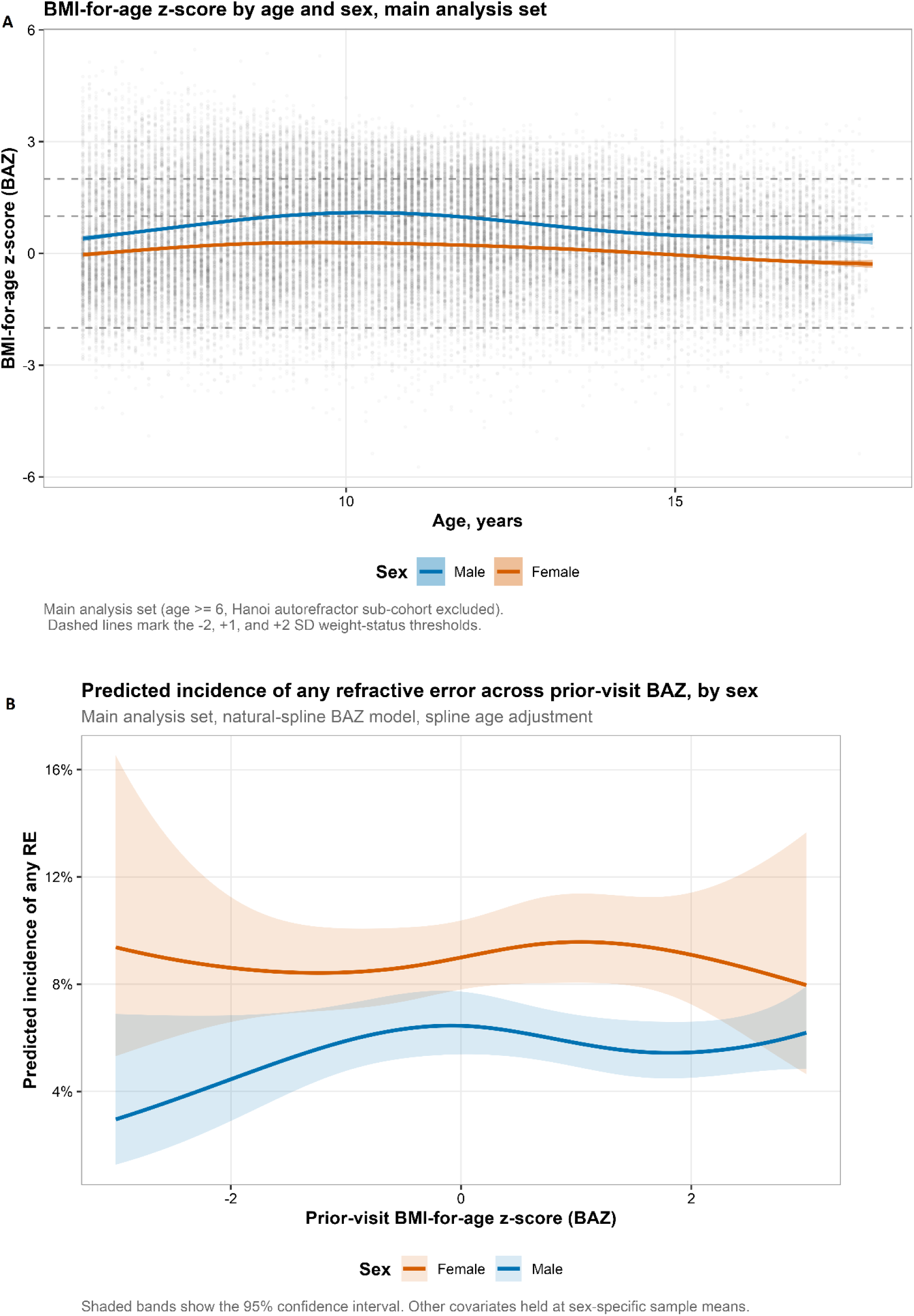
BMI-for-age z-score and prevalence of refractive error (RE). A. BMI-for-age z-score by age and sex in the main analysis set. BMI-for-age z-score (BAZ) plotted against age, shown separately for boys (blue) and girls (orange). Individual examinations are shown as lightly shaded points, with a smoothed trend line and its 95% confidence band overlaid for each sex. Horizontal dashed lines mark the -2, +1, and +2 standard deviation thresholds used to define thinness, overweight, and obesity in children older than 5 years. B. Predicted prevalence of any refractive error across BMI-for-age z-score, by sex. Predicted prevalence of any refractive error across the observed range of BAZ, generated from the natural spline BAZ model adjusted for age (natural spline), city, and examination year, fit separately for boys (blue) and girls (orange) in the main analysis set. Shaded bands show 95% confidence intervals. Other covariates are held at sex-specific sample means. The curve is U shaped in both sexes, lowest near an average BAZ and rising toward both the thin and the obese ends of the distribution, corresponding to the significant nonlinearity detected by a joint Wald test of the spline terms in both boys (p = 0.021) and girls (p = 0.009).

**Table 1.** Characteristics of study examinations according to BMI-for-age category, main analysis set, by sex.

| Characteristic | Male |  |  |  |  |  | Female |  |  |  |  |  |
| --- | --- | --- | --- | --- | --- | --- | --- | --- | --- | --- | --- | --- |
|  | Overall<br>N =<br>25,583<sup>#</sup> | Thin<br>N =<br>672<sup>#</sup> | Normal<br>BMI<br>N =<br>12,856<sup>#</sup> | Overweight<br>N = 6,652<sup>#</sup> | Obesity<br>N =<br>5,403<sup>#</sup> | p-value<sup>*</sup> | Overall<br>N =<br>23,832<sup>#</sup> | Thin<br>N =<br>832<sup>#</sup> | Normal<br>BMI<br>N =<br>17,086<sup>#</sup> | Overweight<br>N = 4,491<sup>#</sup> | Obesity<br>N =<br>1,423<sup>#</sup> | p-value<sup>*</sup> |
| Age, years | 11.0<br>(3.1) | 10.7<br>(3.2) | 11.2<br>(3.4) | 11.4 (2.9) | 10.2<br>(2.5) | <0.001 | 11.2<br>(3.2) | 11.0<br>(3.1) | 11.4<br>(3.3) | 10.8 (2.8) | 10.0<br>(2.7) | <0.001 |
| City |  |  |  |  |  | <0.001 |  |  |  |  |  | <0.001 |
| Hanoi | 16,242<br>(63%) | 466<br>(69%) | 8,341<br>(65%) | 4,229<br>(64%) | 3,206<br>(59%) |  | 14,768<br>(62%) | 550<br>(66%) | 10,793<br>(63%) | 2,620<br>(58%) | 805<br>(57%) |  |
| Ho Chi Minh City | 6,617<br>(26%) | 142<br>(21%) | 3,301<br>(26%) | 1,662<br>(25%) | 1,512<br>(28%) |  | 6,605<br>(28%) | 219<br>(26%) | 4,658<br>(27%) | 1,304<br>(29%) | 424<br>(30%) |  |
| Hai Phong | 2,724<br>(11%) | 64<br>(9.5%) | 1,214<br>(9.4%) | 761 (11%) | 685<br>(13%) |  | 2,459<br>(10%) | 63<br>(7.6%) | 1,635<br>(9.6%) | 567 (13%) | 194<br>(14%) |  |
| Examination year |  |  |  |  |  | <0.001 |  |  |  |  |  | <0.001 |
| 2022 | 1,174<br>(4.6%) | 26<br>(3.9%) | 539<br>(4.2%) | 290<br>(4.4%) | 319<br>(5.9%) |  | 1,056<br>(4.4%) | 23<br>(2.8%) | 693<br>(4.1%) | 248<br>(5.5%) | 92<br>(6.5%) |  |
| 2023 | 12,084<br>(47%) | 341<br>(51%) | 6,096<br>(47%) | 3,139<br>(47%) | 2,508<br>(46%) |  | 11,322<br>(48%) | 404<br>(49%) | 8,128<br>(48%) | 2,122<br>(47%) | 668<br>(47%) |  |
| 2024 | 12,325<br>(48%) | 305<br>(45%) | 6,221<br>(48%) | 3,223<br>(48%) | 2,576<br>(48%) |  | 11,454<br>(48%) | 405<br>(49%) | 8,265<br>(48%) | 2,121<br>(47%) | 663<br>(47%) |  |

#Mean (SD); n (%)
\*One-way analysis of means; Pearson's Chi-squared test

Children with obesity were on average about a year younger than children with normal BMI in both sexes (10.2 versus 11.2 years in boys, 10.0 versus 11.4 years in girls), and BMI status differed by city and examination year in both sexes (all p<0.001) (**Table 1**).

### Refractive error according to BMI-for-age category

In the main analysis set, crude prevalence of any refractive error among boys was 49.1% in thinness, 47.6% in normal weight, 51.6% in overweight, and 47.2% in obesity (**Table 2**). Among girls, the corresponding values were 55.3%, 57.4%, 56.5%, and 53.0%. These crude patterns did not rise monotonically with weight status. Obese children in this study are systematically younger, and refractive error becomes more common as children move through school, so crude numbers understate any true association with adiposity.

**Table 2.** Crude prevalence of refractive errors according to BMI-for-age category, by sex.

| Outcome | Sex | N<br>Thinness | N<br>Normal<br>BMI | N<br>Overweight | N<br>Obesity | Prevalence,<br>% (95%<br>CI)<br>Thinness | Prevalence,<br>% (95% CI)<br>Normal BMI | Prevalence,<br>% (95% CI)<br>Overweight | Prevalence,<br>% (95%<br>CI)<br>Obesity |
| --- | --- | --- | --- | --- | --- | --- | --- | --- | --- |
| Any RE (main analysis set) | Female | 832 | 17,086 | 4,491 | 1,423 | 55.3 (51.9 to 58.6) | 57.4 (56.6 to 58.1) | 56.5 (55.0 to 57.9) | 53.0 (50.4 to 55.6) |
|  | Male | 672 | 12,856 | 6,652 | 5,403 | 49.1 (45.3 to 52.9) | 47.6 (46.8 to 48.5) | 51.6 (50.4 to 52.8) | 47.2 (45.9 to 48.6) |
| Myopia (Hanoi sub-cohort) | Female | 49 | 5,430 | 442 | 160 | 4.1 (1.1 to 13.7) | 6.9 (6.3 to 7.6) | 6.8 (4.8 to 9.5) | 6.9 (3.9 to 11.9) |
|  | Male | 65 | 5,745 | 680 | 488 | 9.2 (4.3 to 18.7) | 6.8 (6.1 to 7.5) | 8.4 (6.5 to 10.7) | 7.6 (5.6 to 10.3) |
| Moderate/high myopia (Hanoi sub-cohort) | Female | 49 | 5,430 | 442 | 160 | 0.0 (0.0 to 7.3) | 0.4 (0.3 to 0.7) | 0.7 (0.2 to 2.0) | 1.9 (0.6 to 5.4) |
|  | Male | 65 | 5,745 | 680 | 488 | 0.0 (0.0 to 5.6) | 0.5 (0.4 to 0.7) | 0.7 (0.3 to 1.7) | 0.8 (0.3 to 2.1) |
| Hyperopia (Hanoi sub-cohort) | Female | 49 | 5,430 | 442 | 160 | 77.6 (64.1 to 87.0) | 71.8 (70.6 to 73.0) | 75.1 (70.9 to 78.9) | 72.5 (65.1 to 78.8) |
|  | Male | 65 | 5,745 | 680 | 488 | 70.8 (58.8 to 80.4) | 71.8 (70.6 to 73.0) | 75.3 (71.9 to 78.4) | 72.3 (68.2 to 76.1) |
| Astigmatism (Hanoi sub-cohort) | Female | 49 | 5,430 | 442 | 160 | 67.3 (53.4 to 78.8) | 65.4 (64.1 to 66.6) | 70.6 (66.2 to 74.6) | 66.9 (59.3 to 73.7) |
|  | Male | 65 | 5,745 | 680 | 488 | 64.6 (52.5 to 75.1) | 67.6 (66.3 to 68.8) | 72.5 (69.0 to 75.7) | 75.8 (71.8 to 79.4) |
| Anisometropia (Hanoi sub-cohort) | Female | 49 | 5,430 | 442 | 160 | 12.2 (5.7 to 24.2) | 17.8 (16.8 to 18.8) | 17.0 (13.8 to 20.7) | 14.4 (9.8 to 20.6) |
|  | Male | 65 | 5,745 | 680 | 488 | 15.4 (8.6 to 26.1) | 16.5 (15.6 to 17.5) | 18.8 (16.1 to 21.9) | 21.5 (18.1 to 25.4) |

Any refractive error (RE) is estimated in the main analysis set (age $\geq 6$ , Hanoi autorefractor sub-cohort excluded), n = 25583 male, 23832 female. Myopia, moderate/high myopia, hyperopia, astigmatism, and anisometropia are estimated in the independent Hanoi autorefractor sub-cohort, n = 6978 male, 6081 female. 95% CI: 95% confidence interval.

After adjustment for age (natural spline), city, and examination year, each 1-SD increase in BAZ was associated with a small increase in any refractive error prevalence in girls (adjusted prevalence ratio [aPR] 1.01, 95% CI 1.00 to 1.02, p=0.027), while the association in boys was weaker and not significant (aPR 1.01, 95% CI 1.00 to 1.02, p=0.099) (**Table 3**). A joint Wald test of the spline terms confirmed significant nonlinearity in both boys (p=0.021) and girls (p=0.009) (**Table S4**), and the adjusted curves were U shaped in both sexes, lowest near a BAZ close to zero and rising again toward both thinness and obesity (**Figure 1**).

**Table 3.**
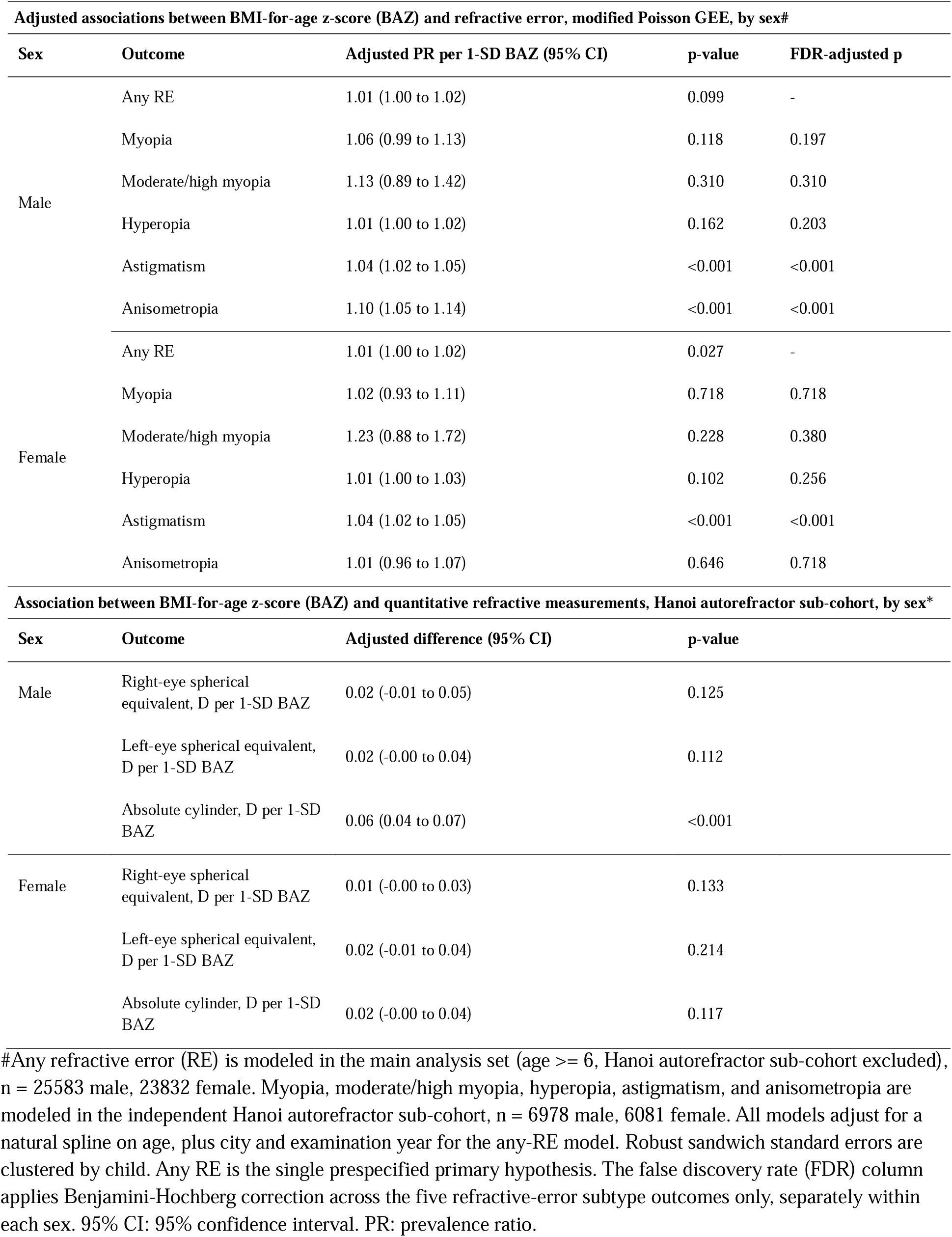
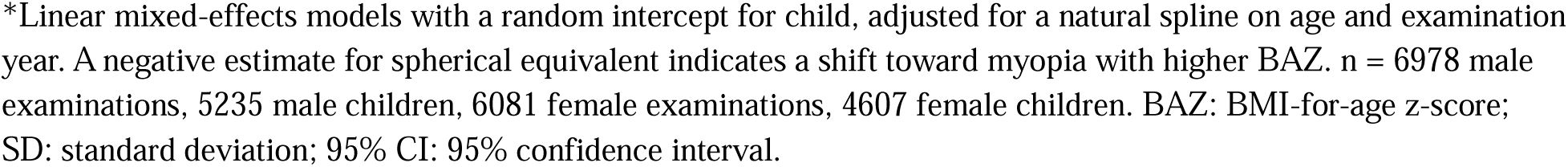
Adjusted associations between BMI-for-age z-score (BAZ) and refractive error, and quantitative refractive measurements, by sex.

| Adjusted associations between BMI-for-age z-score (BAZ) and refractive error, modified Poisson GEE, by sex# |  |  |  |  |
| --- | --- | --- | --- | --- |
| Sex | Outcome | Adjusted PR per 1-SD BAZ (95% CI) | p-value | FDR-adjusted p |
| Male | Any RE | 1.01 (1.00 to 1.02) | 0.099 | - |
|  | Myopia | 1.06 (0.99 to 1.13) | 0.118 | 0.197 |
|  | Moderate/high myopia | 1.13 (0.89 to 1.42) | 0.310 | 0.310 |
|  | Hyperopia | 1.01 (1.00 to 1.02) | 0.162 | 0.203 |
|  | Astigmatism | 1.04 (1.02 to 1.05) | <0.001 | <0.001 |
|  | Anisometropia | 1.10 (1.05 to 1.14) | <0.001 | <0.001 |
| Female | Any RE | 1.01 (1.00 to 1.02) | 0.027 | - |
|  | Myopia | 1.02 (0.93 to 1.11) | 0.718 | 0.718 |
|  | Moderate/high myopia | 1.23 (0.88 to 1.72) | 0.228 | 0.380 |
|  | Hyperopia | 1.01 (1.00 to 1.03) | 0.102 | 0.256 |
|  | Astigmatism | 1.04 (1.02 to 1.05) | <0.001 | <0.001 |
|  | Anisometropia | 1.01 (0.96 to 1.07) | 0.646 | 0.718 |
| Association between BMI-for-age z-score (BAZ) and quantitative refractive measurements, Hanoi autorefractor sub-cohort, by sex* |  |  |  |  |
| Sex | Outcome | Adjusted difference (95% CI) | p-value |  |
| Male | Right-eye spherical equivalent, D per 1-SD BAZ | 0.02 (-0.01 to 0.05) | 0.125 |  |
|  | Left-eye spherical equivalent, D per 1-SD BAZ | 0.02 (-0.00 to 0.04) | 0.112 |  |
|  | Absolute cylinder, D per 1-SD BAZ | 0.06 (0.04 to 0.07) | <0.001 |  |
| Female | Right-eye spherical equivalent, D per 1-SD BAZ | 0.01 (-0.00 to 0.03) | 0.133 |  |
|  | Left-eye spherical equivalent, D per 1-SD BAZ | 0.02 (-0.01 to 0.04) | 0.214 |  |
|  | Absolute cylinder, D per 1-SD BAZ | 0.02 (-0.00 to 0.04) | 0.117 |  |

**Table S1** breaks the BAZ association down by weight status category, with normal weight as reference, consistent with this nonlinear pattern. Obesity was associated with higher any refractive error prevalence in girls (aPR 1.05, 95% CI 1.01 to 1.10, p=0.017), with a similar but nonsignificant estimate in boys (aPR 1.03, 95% CI 1.00 to 1.06, p=0.059). Neither overweight nor thinness reached significance in either sex, though the thinness estimate in boys ran the same direction (1.05, p=0.089). Together with **Figure 1**, this suggests an association concentrated at both tails of the BMI distribution rather than a smooth gradient.

### Specific refractive-error phenotypes and quantitative refractive measurements

In the independent Hanoi autorefractor subgroup (6,978 male and 6,081 female examinations), BAZ was not significantly associated with myopia in either sex (aPR 1.06, 95% CI 0.99 to 1.13, p=0.118 in boys, 1.02, 95% CI 0.93 to 1.11, p=0.718 in girls, **Table 3, Figure S2**), nor with moderate or high myopia (aPR 1.13, 95% CI 0.89 to 1.42 in boys, 1.23, 95% CI 0.88 to 1.72 in girls). Crude prevalence showed similarly modest differences across categories (**Table 2**, **Figure 2**).

**Figure 2.**
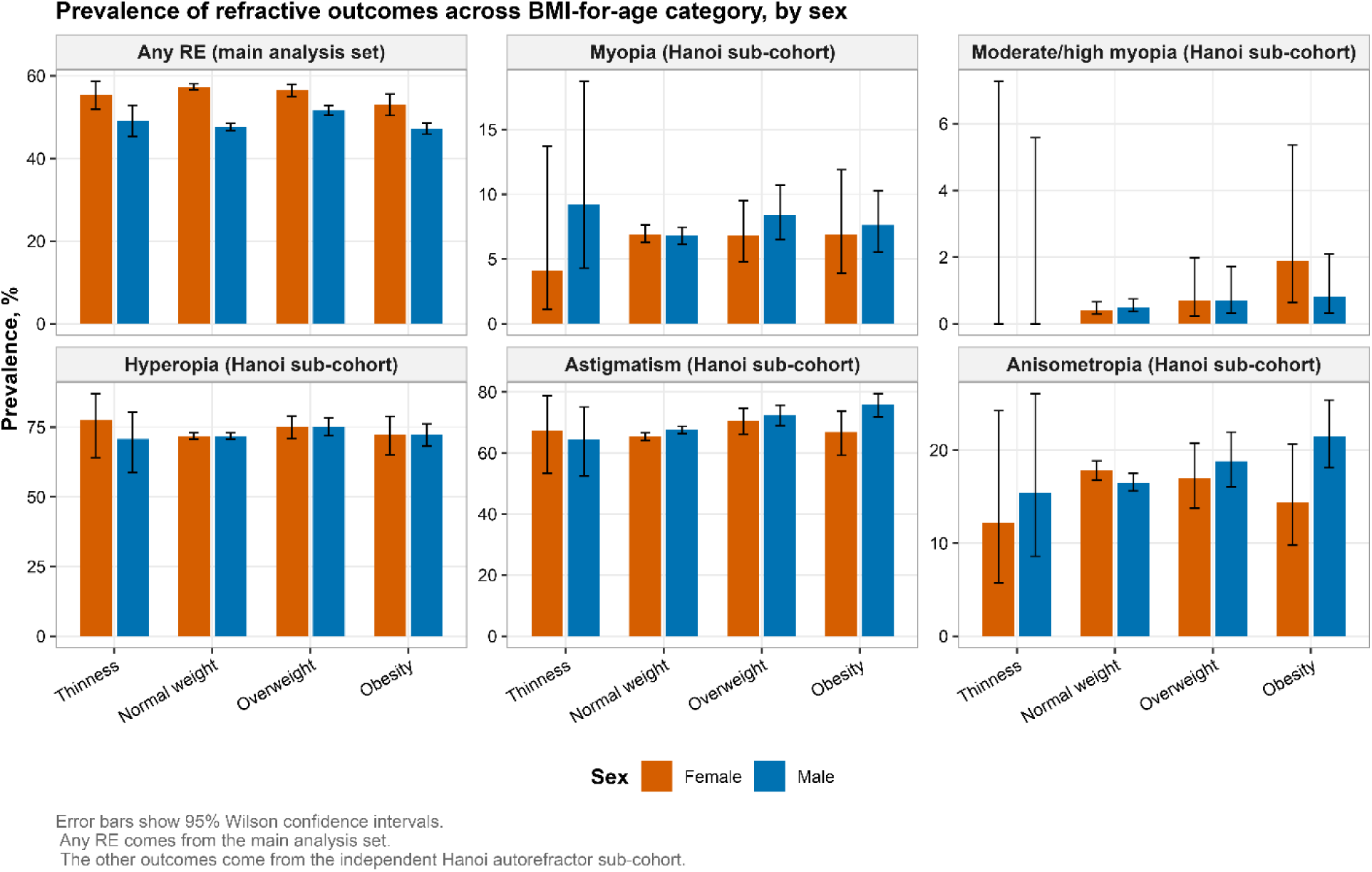
Crude prevalence of refractive outcomes by BMI-for-age category, by sex. Unadjusted prevalence of six refractive outcomes across four BMI-for-age categories, thinness, normal BMI, overweight, and obesity, shown separately for boys (blue) and girls (orange). Any refractive error is estimated in the main analysis set. Myopia, moderate or high myopia, hyperopia, astigmatism, and anisometropia are estimated in the independent Hanoi autorefractor sub-cohort. Error bars show 95% Wilson confidence intervals. Vertical axis scales differ across panels to accommodate the very different baseline prevalence of each outcome, and should not be compared directly across panels.

In contrast, higher BAZ was consistently associated with astigmatism, with a 4% higher prevalence per 1-SD BAZ in both boys and girls (aPR 1.04, 95% CI 1.02 to 1.05, p<0.001 for both), significant after false discovery rate adjustment (**Table 3**). Anisometropia was associated with higher BAZ in boys (aPR 1.10, 95% CI 1.05 to 1.14, p<0.001) but not girls (aPR 1.01, 95% CI 0.96 to 1.07, p=0.646). Hyperopia was not associated with BAZ in either sex.

Quantitative analyses provided further evidence against a myopic shift with adiposity (**Table 3, Figure S3**). Each 1-SD increase in BAZ was associated with a 0.02-D higher right-eye spherical equivalent in boys (95% CI -0.01 to 0.05, p=0.125) and a similarly small, nonsignificant change in girls, with left-eye spherical equivalent showing the same pattern. Absolute cylinder increased significantly with BAZ in boys (0.06 D, 95% CI 0.04 to 0.07, p<0.001) but not girls (0.02 D, 95% CI -0.00 to 0.04, p=0.117).

### Heterogeneity between cities and sensitivity analyses

The association between BAZ and any refractive error varied by city (**Figure S1, Table S2**). Among girls, the interaction with Ho Chi Minh City was significant (p=0.005) but not with Hai Phong (p=0.750). Among boys, neither city interaction was significant (Ho Chi Minh City p=0.146, Hai Phong p=0.788). City-specific analyses (**Table S3**) showed the predicted curve rising mainly in Ho Chi Minh City for boys (aPR 1.05, 95% CI 1.03 to 1.08, p<0.001) and mainly in Hanoi for girls (aPR 1.02, 95% CI 1.00 to 1.03, p=0.008), with associations weaker and nonsignificant elsewhere and a flat slope in Hai Phong for both sexes.

Results were broadly similar when analyses included children aged 3 to 18 years, although statistical significance was attenuated among boys (p=0.053) and was borderline among girls (p=0.050). Excluding current spectacle wearers also produced estimates that were directionally similar to the primary analysis (**Table S3**).

### Longitudinal replication check

Prior-visit BAZ did not predict incident any RE in either sex (aPR 1.02 per 1-SD BAZ, 95% CI 0.94 to 1.09, p = 0.681 in boys and 1.02, 95% CI 0.94 to 1.11, p = 0.619 in girls) (**Table 4**). The nonlinear, U shaped pattern did not reappear, with no evidence of nonlinearity in either sex (p = 0.310 in boys, p = 0.796 in girls) (**Figure S4**). A second check using each child’s mean BAZ across their full observation window to predict any RE by their last visit gave the same null result (aPR 1.03, p=0.450 in boys, 1.02, p=0.709 in girls). Several candidate refractive error subtypes could not be meaningfully tested due to small numbers.

**Table 4.** Longitudinal replication check summary.

| **Adjusted associations between prior-visit BAZ and incident refractive error, by sex#** |  |  |  |
| --- | --- | --- | --- |
| Sex | Outcome | Adjusted PR per 1-SD BAZ (95% CI) | p-value |
| Male | Any RE | 1.02 (0.94 to 1.09) | 0.681 |
| Female | Any RE | 1.02 (0.94 to 1.11) | 0.619 |
| **Test for nonlinearity in the BAZ to incident any-RE association, by sex** |  |  |  |
| Sex | Test |  | p-value |
| Male | Joint Wald test, BAZ natural-spline terms |  | 0.310 |
| Female | Joint Wald test, BAZ natural-spline terms |  | 0.796 |
| **Cumulative (mean) BAZ across all visits and incident any RE by the child's last visit, by sex\$** | | | |
| Sex | Children analyzed | Adjusted PR per 1-SD mean BAZ (95% CI) | p-value |
| Male | 4,384 | 1.03 (0.95 to 1.11) | 0.450 |
| Female | 3,286 | 1.02 (0.93 to 1.11) | 0.709 |

## DISCUSSION

In this large, multi-city Vietnamese data, adiposity was not simply linked to more refractive error. The association was nonlinear, sex specific, and concentrated in particular subtypes rather than myopia itself, following a U shaped curve across BAZ that was lowest near an average BMI and higher at both extremes, significant in girls but only bordering on it in boys. Obesity was linked to modestly higher any refractive error prevalence in girls, astigmatism to BAZ in both sexes, and anisometropia and higher cylinder only in boys, while myopia, moderate or high myopia, and hyperopia showed no clear association, and spherical equivalent shifted slightly toward hyperopia rather than myopia.

The U shaped pattern echoes the nationwide Israeli study of 1.36 million adolescents that also found both underweight and high BMI associated with myopia, with a stronger effect in males ^5^. It does not match the inverse L shaped curve reported in a smaller Chinese schoolchildren sample ^8^, suggesting the shape may depend on age range and how finely BMI is modeled. Our finding of significance mainly in girls fits Korean and Chinese data showing obesity carrying a 3.77 fold higher odds of high myopia in girls ^6^, and where overweight and obesity were more strongly tied to myopia in girls age 6 to 11 ^7^.

Where our results diverge more clearly from prior work is in outcome specificity. Much of the existing literature, including a 2025 meta-analysis reporting pooled odds ratios of 1.27 for overweight and 1.25 for obesity ^4^, NHANES analyses in American adolescents ^10^, and the Korean cohorts already cited ^6,7^, frames the adiposity association mainly around myopia. We found no significant myopia association in either sex, while astigmatism and, in boys, anisometropia carried the clearest signal. This lines up with a pediatric study linking obesity to roughly double the odds of astigmatism ^9^, and with a recent narrative review concluding that BMI ties more consistently to astigmatism than to hyperopia, while flagging hyperopia evidence as inconclusive ^22^. The modest hyperopic, rather than myopic, shift we observed is unexpected and needs replication.

The BAZ by city interaction, strongest for Ho Chi Minh City in boys and for Hanoi in girls, is a reminder that this association is not likely to be biologically fixed. Near work, outdoor time, screening practices, and schooling patterns differ across Vietnamese cities and were not measured here, and any could modify or confound the BAZ association, a concern already raised in literature generally ^6,12,22^. A Mendelian randomization analysis that failed to support a causal genetic link between BMI and myopia reinforces the same caution ^11^.

A supplementary longitudinal check using consecutive visit pairs found no evidence that prior BAZ predicts the later onset of refractive error, and the nonlinear pattern central to our main cross-sectional finding did not reappear. This argues that the cross-sectional association may reflect confounding or reverse relationships rather than adiposity driving new refractive change. We are cautious about our RE incidence analysis, however, because almost all of the available visit pairs were only one year apart, with very few spanning two years, and refractive error is generally understood to develop gradually over several years rather than within a single annual cycle. A one-year window may simply be too short to detect a genuine effect of adiposity on the development of refractive error, especially if the underlying mechanism, whether behavioral or metabolic, operates on a longer time scale.

This study has noteworthy limitations. Refractive error subtypes and quantitative measurements came only from the Hanoi autorefractor sub-cohort, so those findings may not generalize to the other cities. Hyperopia was defined broadly enough to capture physiological hyperopia common in young children, which may have diluted any true signal. Because our analysis is mainly cross-sectional, and the short-follow-up check was mostly null, we have not established whether adiposity precedes refractive change, both share a common driver, or early refractive error changes a child’s activity and weight over time. Thus, longitudinal analysis with more years of data would be the natural next step.

Taken together, these findings suggest that in Vietnamese schoolchildren, obesity carries a small but significant association with refractive error that differs by sex and by city. For school health practice, this argues for watching astigmatism and anisometropia, not only myopia, and for not assuming only children with obesity are at risk, given the rise seen at low BAZ as well.

## DECLARATIONS

### Ethical Statement

The study was approved by the Vinmec Ethics Committee (approval number 0231/2024/CN/HDDD VMEC). Written informed consent was waived for secondary analysis of de-identified, retrospective data.

### Author contribution

N.T.H. did conceptualization, data curation, formal analysis, investigation, methodology, project administration, resources, software, supervision, validation, visualization, writing original draft, and writing review & editing.

K.P.M.T. did data cleaning and writing review & editing.

All authors read and approved the manuscript.

### Data Availability Statement

R code is available from the corresponding author upon reasonable request. Individual patient-level data cannot be shared due to applicable privacy regulations and the terms of the institutional ethics approval.

### Funding source

This study did not receive any funding.

### Declaration of Interests

None of the authors have any proprietary interests or conflicts of interest related to this submission.

### Declaration of generative AI use

All scientific content, analyses, and interpretations are the original work of the authors. The authors used AI-assisted tools (Claude Sonet 5) for language editing and grammar checking during manuscript preparation. Originality and accuracy of content has been confirmed by the authors. The authors have checked terms of use for the specific AI tool used, and therefore confirm suitability for publication. The authors take full responsibility for the integrity of the whole content, including accuracy of references.

## Notes

### Competing Interest Statement

The authors have declared no competing interest.

