## Supplementary Tables and Figures for "Adiposity and the Prevalence, Incidence and Distribution of Refractive Errors in Urban Vietnamese Children"

SUPPLEMENTARY FIGURES AND TABLES

Figure S1. Predicted prevalence of any refractive error across BMI-for-age z-score, by city and sex.
Predicted prevalence of any refractive error across BAZ from city-specific modified Poisson models, fit separately for boys and girls within each of the three study cities in the main analysis set. Shaded bands show 95% confidence intervals. The association with BAZ is steepest for boys in Ho Chi Minh City and for girls in Hanoi, while the slope in Hai Phong is close to flat for both sexes, a pattern consistent with the significant BAZ by city interaction detected for Ho Chi Minh City in the interaction testing.

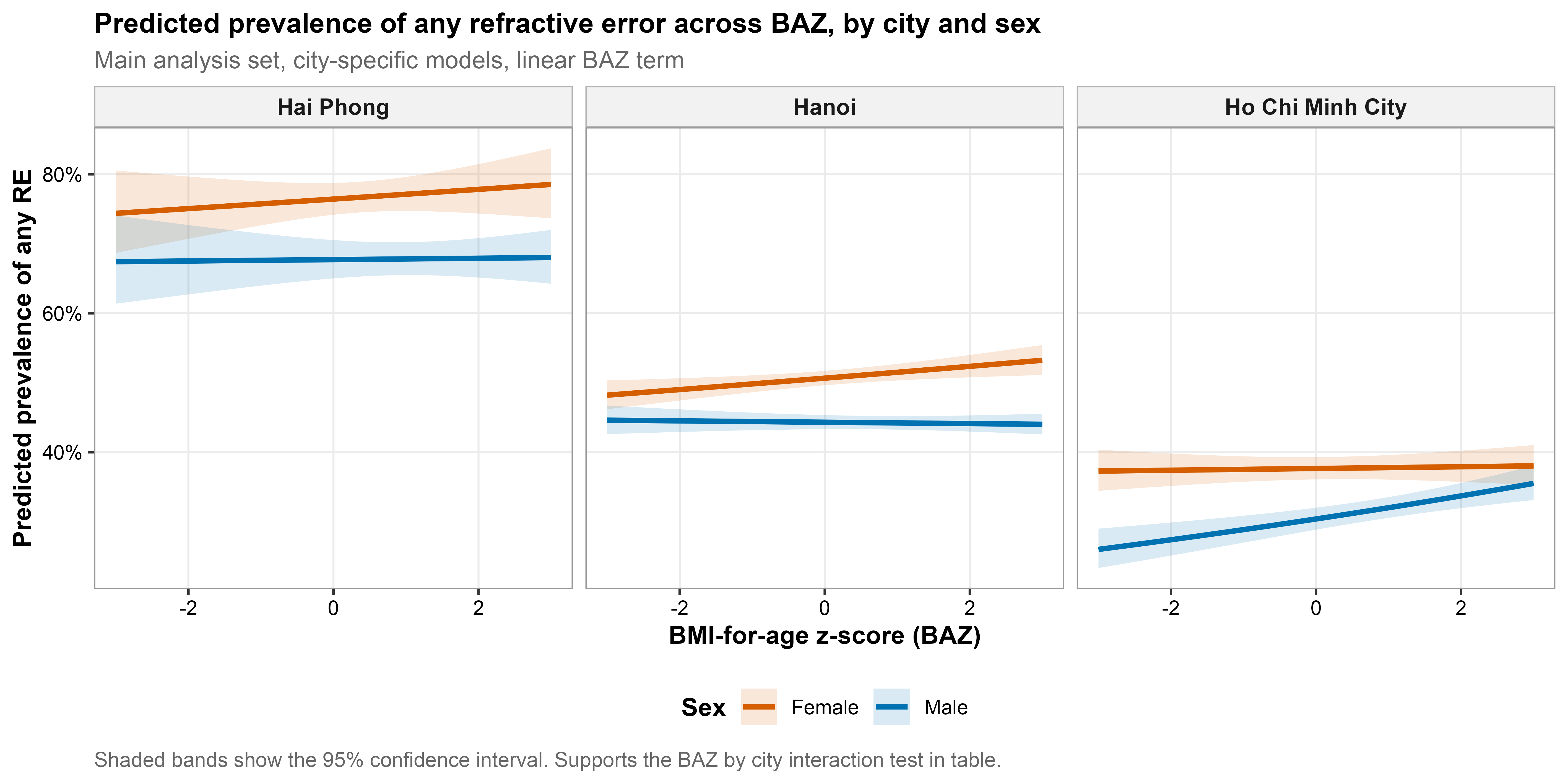

Figure S2. Predicted prevalence of myopia across BMI-for-age z-score, by sex.
Predicted prevalence of myopia across BAZ from the adjusted modified Poisson model, fit with generalized estimating equations, natural spline age adjustment, and examination year, fit separately for boys and girls in the independent Hanoi autorefractor sub-cohort. Shaded bands show 95% confidence intervals. The curve is nearly flat in both sexes, consistent with the absence of a statistically significant association between BAZ and myopia after adjustment.

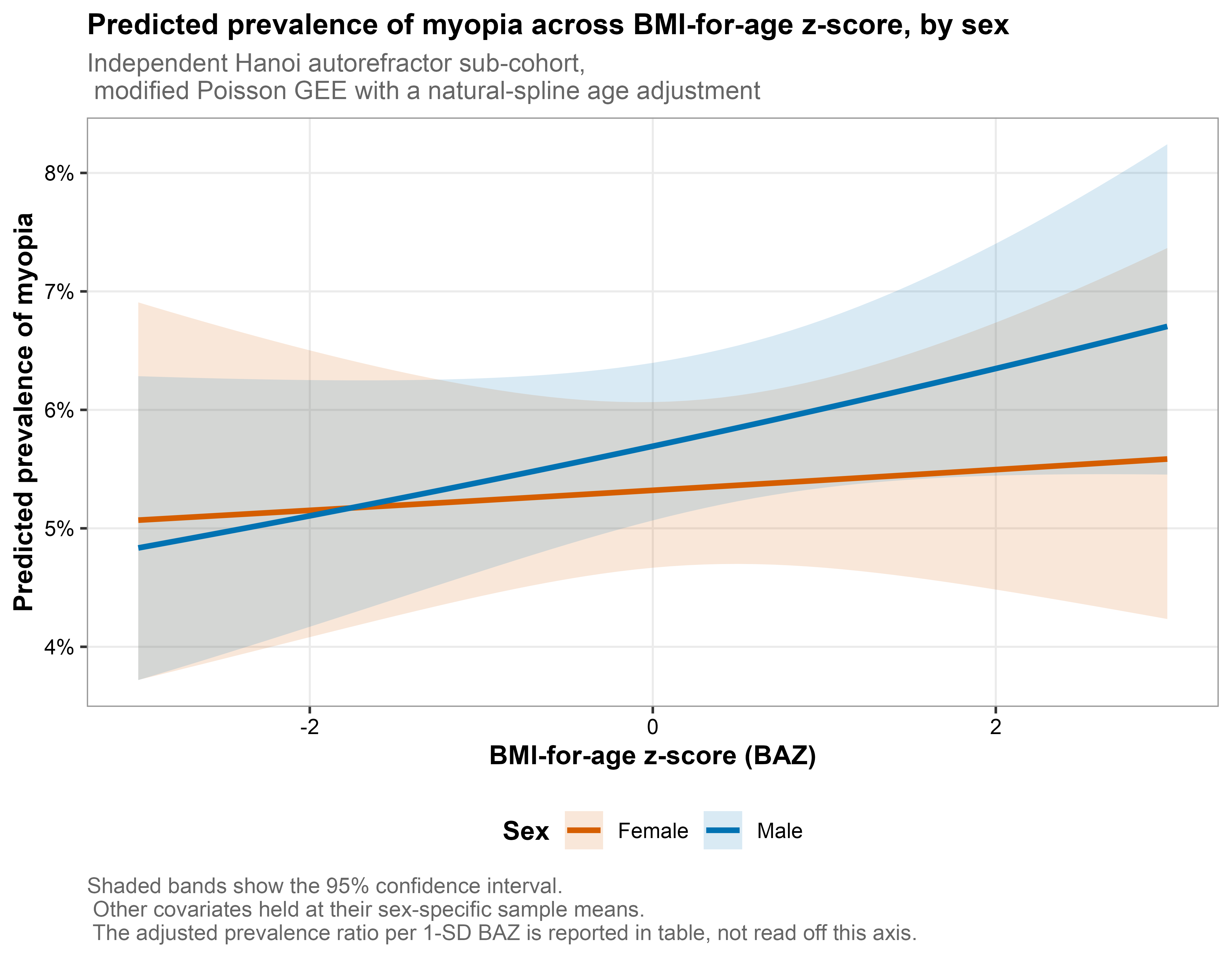

Figure S3. Adjusted right-eye spherical equivalent across BMI-for-age z-score, by sex.
Predicted right-eye spherical equivalent, in diopters, across BAZ from a linear mixed-effects model with natural splines on BAZ and age and a random intercept for child, fit separately for boys and girls in the independent Hanoi autorefractor sub-cohort. Shaded bands show 95% confidence intervals. The horizontal dashed line marks 0 diopters, or emmetropia. Lower, more negative values indicate a more myopic refractive state. Spherical equivalent rises gently with increasing BAZ in both sexes, indicating a shift toward hyperopia rather than myopia at higher adiposity.

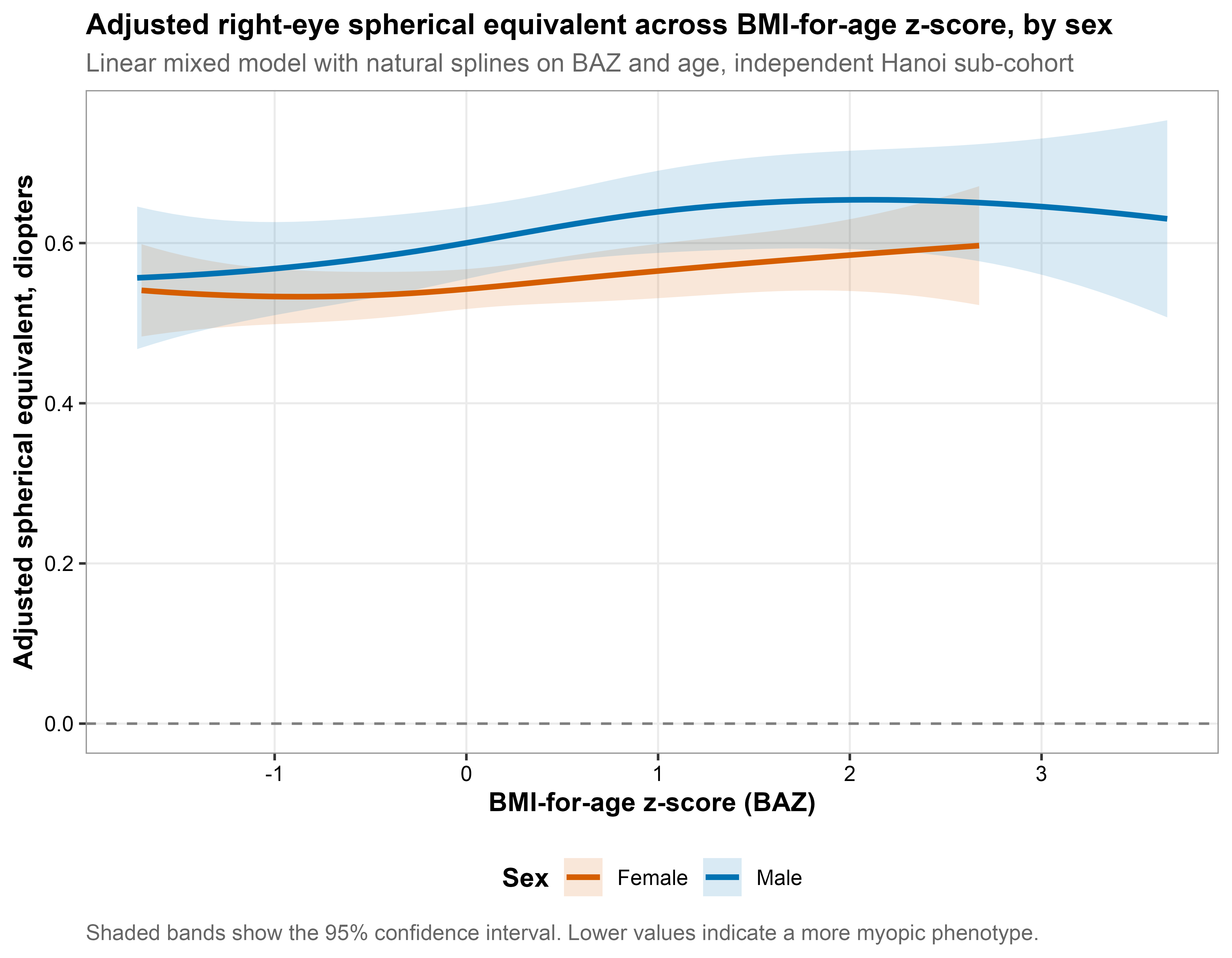

Figure S4. Predicted incidence of any refractive error across prior-visit BMI-for-age z-score, by sex.
Predicted incidence of new-onset any refractive error across BAZ measured at the earlier visit in a pair of consecutive annual examinations, fit separately for boys and girls among children free of any refractive error at the earlier visit, adjusted for age at the later visit (natural spline), city, and examination year. Shaded bands show 95 percent confidence intervals. Unlike the cross-sectional U shaped curve in Figure 3, this curve is essentially flat in both sexes, consistent with the null prevalence ratio reported for this supplementary longitudinal check (1.02 in both sexes, p greater than 0.6). Nearly all visit pairs contributing to this figure were 1 year apart, so a flat curve here should be read as inconclusive over a short follow-up interval, not as evidence against an effect over a longer one.

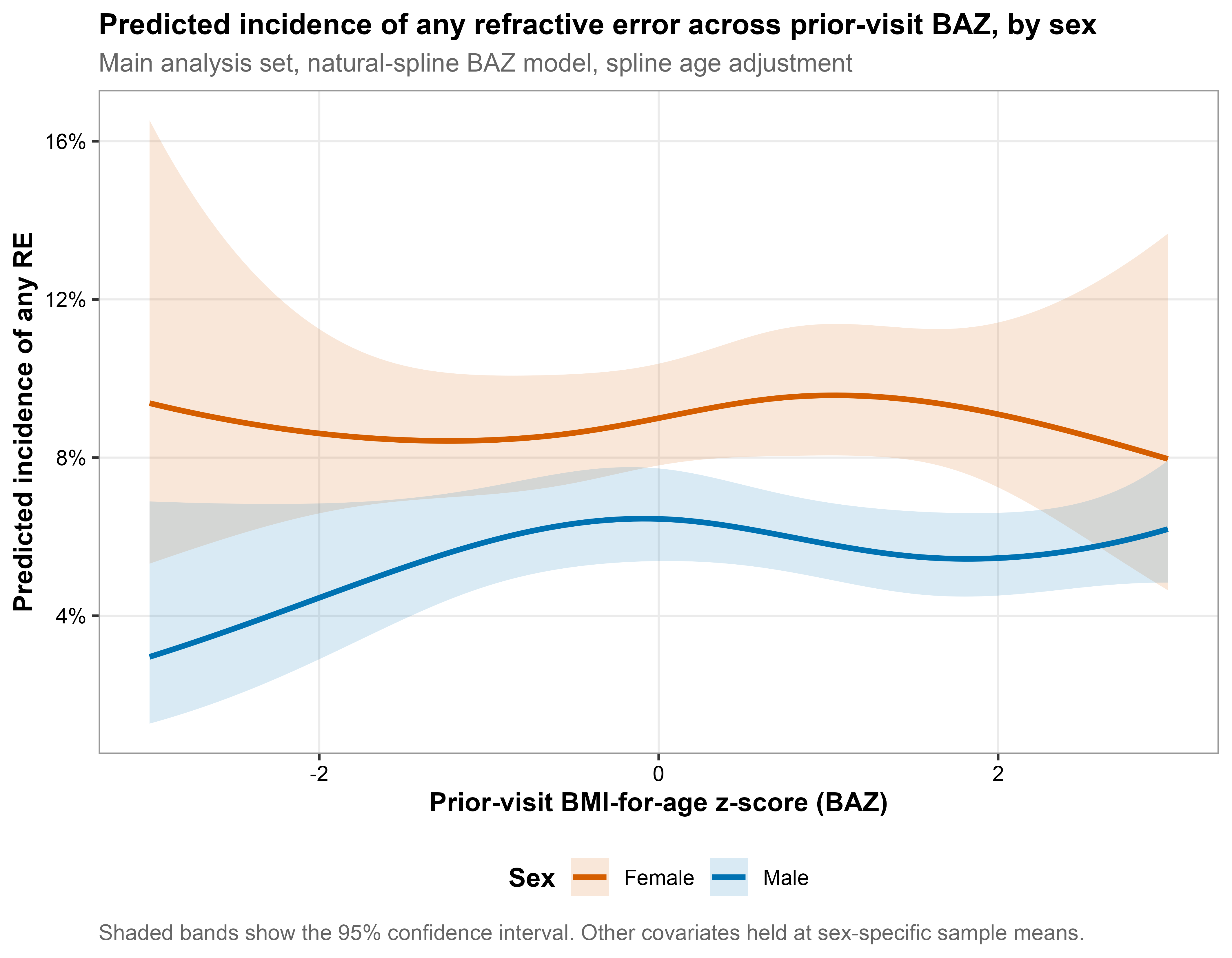

Table S1. Association between weight-status category and any refractive error, by sex.

| Sex | Term | aPR (95% CI) | p-value |
| --- | --- | --- | --- |
| Male | Thinness | 1.05 (0.99 to 1.10) | 0.089 |
|  | Overweight | 1.02 (1.00 to 1.04) | 0.098 |
|  | Obesity | 1.03 (1.00 to 1.06) | 0.059 |
| Female | Thinness | 1.00 (0.95 to 1.04) | 0.886 |
|  | Overweight | 1.02 (0.99 to 1.05) | 0.127 |
|  | Obesity | 1.05 (1.01 to 1.10) | 0.017 |
| Reference category: normal weight. Adjusted for a natural spline on age, city, and examination year. Main analysis set only. Obesity is associated with any RE in females and borderline in males. While overweight and thinness are not distinguishable from normal weight. | | | |

Table S2. Prespecified interaction test for the BAZ to any-RE association, by sex.

| Sex | Interaction | p-value |
| --- | --- | --- |
| Male | BAZ x city (Ho Chi Minh City) | 0.146 |
|  | BAZ x city (Hai Phong) | 0.788 |
| Female | BAZ x city (Ho Chi Minh City) | 0.005 |
|  | BAZ x city (Hai Phong) | 0.750 |
| Exploratory, fitted in publication run mode only. A model shown as not run reflects RUN_MODE, not a failed fit. | | |

Table S3. Sensitivity analyses for the BAZ to any-RE association, by sex.

| Sensitivity_analysis | Adjusted PR (95% CI) | p-value |
| --- | --- | --- |
| Male, primary model | 1.01 (1.00 to 1.02) | 0.099 |
| Male, raw BMI (BMI term shown for reference only) | 1.00 (1.00 to 1.01) | 0.168 |
| Male, Hanoi only (non-autorefractor examinations) | 1.00 (0.99 to 1.01) | 0.695 |
| Male, Ho Chi Minh City only | 1.05 (1.03 to 1.08) | <0.001 |
| Male, Hai Phong only | 1.00 (0.98 to 1.02) | 0.897 |
| Male, excluding current spectacle wearers | 0.98 (0.95 to 1.02) | 0.390 |
| Male, age 3-18 (no age floor) | 1.01 (1.00 to 1.02) | 0.053 |
| Female, primary model | 1.01 (1.00 to 1.02) | 0.027 |
| Female, raw BMI (BMI term shown for reference only) | 1.00 (1.00 to 1.01) | 0.023 |
| Female, Hanoi only (non-autorefractor examinations) | 1.02 (1.00 to 1.03) | 0.008 |
| Female, Ho Chi Minh City only | 1.00 (0.98 to 1.03) | 0.763 |
| Female, Hai Phong only | 1.01 (0.99 to 1.03) | 0.418 |
| Female, excluding current spectacle wearers | 1.02 (0.98 to 1.06) | 0.309 |
| Female, age 3-18 (no age floor) | 1.01 (1.00 to 1.02) | 0.050 |
| Unless noted otherwise, rows use the main analysis set (age >= 6, Hanoi autorefractor sub-cohort excluded) and a natural-spline age adjustment. The raw BMI row reports the coefficient on raw BMI rather than BAZ and is not on the same scale as the other rows. The age 3-18 row drops the age floor while keeping the same Hanoi autorefractor exclusion, to show whether that restriction changes the qualitative result rather than just sharpening it. | | |

Table S4. Test for nonlinearity in the BAZ to any-RE association, by sex.

| Sex | Test | Statistic | df | p-value |
| --- | --- | --- | --- | --- |
| Male | Joint Wald test, BAZ natural-spline terms (Model 4) | 9.68 | 3 | 0.021 |
| Female | Joint Wald test, BAZ natural-spline terms (Model 4) | 11.67 | 3 | 0.009 |
| A small p-value argues against a simple linear BAZ term and supports the spline-based Figure 3 curve over a single adjusted PR. | | | | |
